# Brief: Implementation of a Novel Clinic/Community Partnership Addressing Food Insecurity Among Adults with HIV in the Southern United States

**DOI:** 10.1101/2022.04.29.22274433

**Authors:** Wesli H. Turner, Emma Sophia Kay, James L. Raper, Karen Musgrove, Kathy Gaddis, Anastasia Ferrell, Donna Yester, Joshua Glenn, Meredith Atwater, Kris Hutchins, Ashutosh Tamhane, Dustin M. Long, Polly Kellar, Tom Creger, Ellen Eaton, Amanda L. Willig

**Author notes:** Corresponding Author: Amanda L. Willig. Author Contributions* Wesli H. Turner conducted interviews with program staff as part of the evaluation team and drafted the manuscript. Emma Sophia Kay oversaw program development and collated information for the paper at Birmingham AIDS Outreach (BAO) and edited the manuscript. James L. Raper oversees all 340B-related funded activities and helped develop the program, and provided critical edits to the manuscript. Karen Musgrove is responsible for overall oversight of programs at BAO and drafted the manuscript. Kathy Gaddis led development and initiation of the social worker and nursing transitions for the program and provided critical feedback in the timeline for the manuscript. Anastasia Ferrell oversees nutritional programs at BAO and provided critical feedback in developing and revising the manuscript. Donna Yester developed and implemented nutrition services for the program and provided critical revisions for the manuscript. Joshua Glenn oversees educational programs and meal delivery at BAO and provided critical feedback in developing and revising the manuscript. Meredith Atwater developed and implemented nutrition services for the program and provided critical revisions for the manuscript. Kris Hutchins supervises operations of the food assistance program and provided critical edits to the timeline and content of the manuscript. Ashutosh Tamhane and Dustin Long serve with the 340B evaluation team, assisted with data collation and presentation, and provided edits to the manuscript. Polly Kellar assisted with financial and programmatic infrastructure for the program and provided critical data and edits for the manuscript. Tom Creger and Ellen Eaton lead the evaluation team and assisted with developing structure of the brief and revision of the manuscript. Amanda L. Willig oversees nutrition-related 340B activities for the 1917 Clinic and drafted the manuscript.

## Abstract

Food insecurity is highly prevalent among people with HIV. Traditional calorie-rich, nutrient poor food assistance programs may improve food security but increase risk for other chronic diseases. This case study describes the process evaluation of a novel clinic/community partnership to provide nutritionally adequate, tailored food assistance to adults with HIV in Alabama. Methods used include semi-structured interviews with program staff at Birmingham AIDS Outreach and the University of Alabama at Birmingham’s 1917 HIV/AIDS Clinic, and analysis of descriptive characteristics of individuals enrolled in the food program for a minimum of one year between 2017-2019. The new program served 1,311 patients and enabled more than 300 previously lost-to-follow-up patients to re-engage in HIV care. The program implementation reviewed here can serve as a roadmap to develop clinic/community partnerships focused on a variety of health outcomes and quality of life among food insecure patients.

---

The Southern United States (US) is disproportionately impacted by the dual burdens of HIV infection and food insecurity (Coleman-Jenson et al., 2017). The South accounts for 52% of new HIV diagnoses (Centers for Disease Control and Prevention, 2019; Reif et al., 2017), and an estimated 24-50% of people with HIV (PWH) are food insecure (Willig et al., 2018). Food insecurity, defined as the inability to obtain food in personally or socially acceptable ways (Coleman-Jenson et al., 2019), may negatively impact HIV treatment and ability to manage chronic health needs (Kalichman et al., 2014; Weiser et al., 2009). Food insecurity impacts the entire HIV care continuum from acquisition to viral suppression (Leddy et al., 2020; Singer et al., 2015; Wang et al., 2011; Weiser et al., 2011).. PWH with food insecurity in the US are referred to standard federal or community-led food assistance programs (Anema et al., 2009). However, a recent systematic review of diet quality among food pantry beneficiaries reported diets were low in fruits, vegetables, and dairy products, and provided inadequate amounts of fiber and several micronutrients (Simmet et al., 2017). Participation in food assistance programs thus can reduce food insecurity, but may inadvertently increase chronic disease risk among program clients (Byker Shanks, 2017; Simmet et al., 2017).

To address the burden of food insecurity in HIV, the University of Alabama at Birmingham (UAB) 1917 Clinic and Birmingham AIDS Outreach (BAO) designed a nutrient rich food program tailored to nutritional and social needs of PWH. Here, we outline the development process for a novel, comprehensive food assistance program utilizing Ryan White grant-related income associated with the 340B Drug Pricing Program. The process described can serve as a foundation for developing clinic/community partnerships to provide food assistance with a meaningful impact on clinical HIV care.

## Rationale

In 1992, the US Congress enacted Section 340B of the Public and Service Act, informally referred to as the 340B Drug Pricing Program (340BHealth). The program requires drug manufacturers to provide front-end discounts on outpatient drugs covered by Medicaid and Medicare Part B to qualified clinics/hospitals. Savings are directed to additional services for vulnerable patient populations, which enables qualified clinics to stretch federal resources to provide comprehensive patient care.

Prior to development of the new food assistance program, BAO maintained a basic nutrition assistance program with monthly food boxes and nutritional supplements for PWH who experience food insecurity. While BAO’s traditional program addressed food insecurity, available items were not always consistent with patients’ dietary restrictions or nutrition needs to increase food sufficiency (defined as food required for a healthy lifestyle) (National Research Council, 2006). Additionally, 1917 Clinic nurses were often the initial point of contact when a patient requested nutritional supplements or other assistance. Nurses reported uncertainty in whether the requested assistance was medically appropriate, and struggled to meet these needs in context of their workload. In 2017, BAO agreed to coordinate the development of an expanded nutrition program with the 1917 Clinic, with oversight and approval provided by HRSA.

## Methods

### Setting

The 1917 Clinic, established in Birmingham, Alabama, 1988 during the height of the AIDS crisis, provides comprehensive care for people diagnosed with HIV in Alabama and neighboring states. Onsite services include primary and subspecialty medical care, case management, medical nutrition therapy, mental health, substance abuse treatment and oral health care (University of Alabama at Birmingham, 2020). The 1917 Clinic receives Health and Human Services Administration’s (HRSA) Ryan White HIV/AIDS Part C funds through the National Institutes of Health (NIH) and the 340B Drug Pricing Program. More recently, the 1917 Clinic embraced an innovative approach to address the growing needs of the HIV community it serves. Utilizing grant-related income provided through the HRSA 340B Drug Pricing Program, the clinic is able to offer critical social services to improve ART adherence and viral suppression for PWH in Birmingham, Alabama, and the surrounding area.

Birmingham AIDS Outreach is the longest serving AIDS Service Organization in Alabama, with a history of successful collaboration with the 1917 Clinic on service and outreach programs for PWH and their families. The mission of BAO is “to enhance the quality of life for people living with HIV/AIDS, at-risk, affected individuals, and the LGBTQ community through outreach, age-appropriate prevention education, and supportive services” (Birmingham AIDS Outreach). BAO was incorporated in 1985 as the first nonprofit, community-based organization dedicated to providing HIV/AIDS prevention education and services for persons/families with HIV/AIDS in Alabama. Today, BAO serves over 1,000 PWH, ranging in age from 18 to 80. All programs offered by BAO are free of charge.

Survey and semi-structured interview feedback with 1917 Clinic and BAO staff and patients was obtained to develop and refine the program. Participants identified five initial areas as most likely to alleviate food insecurity among vulnerable PWH: (1) providing foods with more dietary protein and fiber; (2) providing more fresh and frozen produce; (3) expanding the amount and type of nutritional supplements available (e.g. diabetes, renal, and gut intolerance issues); (4) provision of small equipment (such as can openers for canned goods); and (5) meal delivery for PWH unable to cook for themselves, but ineligible (or on a waiting list) for other meal assistance programs. Existing 1917 Clinic/BAO patients selected the program name, the Birmingham AIDS Outreach Food and Education Delivery (B-FED) program. Semi-structured interviews were also conducted with ten participants after the pilot year (Year 1) of B-FED. The UAB Internal Review Board approved program evaluation and quality improvement activities through a Non-Human Subjects Research protocol.

### B-FED Eligibility and Program Components

Consistent with Ryan White and 340B guidelines, B-FED program eligibility requires that individuals be (1) HIV seropositive, and (2) referred by a B-FED program social worker or dietitian. A patient may be referred if there is indication of food insecurity and/or the patient falls below 400% of the federal poverty level. Enrollment for additional BAO services beyond B-FED is optional. Patients are reassessed by BAO for continued eligibility every six months, and must remain engaged in clinical care with at least annual HIV provider visits.

### Referral Process

With B-FED, the 1917 Clinic social workers and registered dietitians (RDs) assess patient food security status and B-FED eligibility and submit referrals to B-FED through a secure shared data system. The referral is processed by the B-FED Education Specialist or BAO’s Director of Client Services to establish an initial appointment. Emergency service referrals are sent directly to the B-FED Education Specialist for processing.

Following program enrollment, the patient is given an appointment day and time to receive services. This provides structure and gives B-FED personnel an estimate of the number of deliveries to prepare each week. Missed visits are promptly rescheduled. “Walk-in” patients who have medical need for nutrition supplements are seen by a BAO case manager. A patient can determine their own frequency of food pick-up (e.g. monthly or as needed). A few select clients have their monthly services delivered to them on a case-by-case basis.

When patients arrive to receive B-FED services, they first meet with a BAO social worker for an average of 15 minutes to discuss any concerns related to their overall medical and social wellbeing (e.g., housing, employment, general health, etc.). The inclusion of case management within the food program increases the face-time individuals have with social workers and provides an opportunity for intervention if needed.

There is an open line of communication between the 1917 Clinic team, the B-FED Education Specialist, and the B-FED Director of Client Services. This is facilitated by use of a secure, HIPAA-compliant shared records system enabling rapid program referrals and communication between sites. B-FED staff provide a monthly list of any patient referrals they were unable to contact, allowing the 1917 Clinic team members to assist in connecting the patient with B-FED. If a B-FED patient falls out care or is not picking up monthly supplements/food boxes, 1917 Clinic staff are notified to attempt to contact the patient and address any medical or social barriers to B-FED participation. Clinic physicians and nurse practitioners also now include food assistance discussions during patient appointments.

### B-FED Services

B-FED addresses food insecurity through four program pathways: food boxes, food vouchers, nutrition supplements, and home delivered meals. These components are reinforced with food preparation items (e.g. can openers) and educational classes.

#### Food Boxes

Monthly food boxes range in size from 50 to 80 pounds based on contents and typically include fresh fruits and vegetables, canned goods, whole grains, fresh dairy, and dense proteins such as tuna. Boxes are designed to provide at least half of the Recommended Dietary Allowance (RDA) for adults for dietary fiber, protein, and all micronutrients (**Table 1**).

**Table 1.** Contents of a Typical B-FED Food Box as Daily Averages.

|  |
| --- |
| • 2 non-starchy vegetables (fresh)<br>• 1 non-starchy vegetable (frozen)<br>• 2 seasonal fresh fruits<br>• 3 ounces chicken, pork loin, OR beef OR 2 eggs<br>• 3 ounces frozen OR canned fish<br>• ½ cup beans (black, red, pinto, or navy)<br>• 1 cup multigrain pasta OR brown rice OR 2 slices whole wheat bread<br>• 1 cup of oatmeal OR high fiber cereal<br>• 1 ½ cups milk<br>• 2 ounces cheese OR 6 ounces yogurt<br>• 1 serving of dessert (can be fruit, nuts, OR other snack food) |

BAO and the 1917 Clinic have also included an emergency food box component to the program for situations of acute food insecurity. An emergency food box contains the same contents as the monthly food box, apart from Ryan White purchased shelf stable items or the food voucher. Typically, upon receiving an emergency food box referral, BAO will make contact with the patient within 24-48 hours if same-day pick-up is not possible. After receiving an emergency food box, the patient has the option of enrolling into the monthly food box program.

#### Food Voucher

Food vouchers are typically included with the monthly food boxes, but may also be provided as a stand-alone program. The food voucher is a disposable pre-paid card associated with a local supermarket chain. A small number of B-FED patients, typically those who lack proper storage capacity for the food box or are not otherwise able to cook, only receive the food voucher. The vouchers are worth $50.00, do not expire, and can be used to purchase hot meals provided by the grocery deli. The vouchers cannot be used for items other than food.

#### Nutrition Supplements

Nutrition supplements are recommended by 1917 Clinic providers for patients who cannot maintain adequate health through food alone. Prior to B-FED implementation, BAO was limited to providing one case (approximately 24 servings) of one standard supplement per month per patient, regardless of medical need. Additionally, supplement requests could be made by any member of the clinic staff, regardless of medical necessity, leading to difficulties with monitoring safety of supplement use (for example, whether someone with poorly controlled blood sugar was using the supplements appropriately). With the B-FED program, 1917 Clinic RDs assess medical need for supplementation, agree on a usage plan with the patient, and send referrals to the B-FED program. If the referral is for a full year of supplement use, a patient is reassessed in-clinic after 12 months by an RD to determine continued supplement use is necessary. Supplement requests are tailored to the amount, type, and length of time that is most appropriate for the patient. Based on medical need, the program can provide six different brands of liquid nutrition supplements in addition to nutrition bars.

#### Home Delivery Meals

Meal delivery support is offered to patients who are acutely or chronically unable to prepare their own meals (e.g., chronic health conditions, mobility issues, recent surgeries, chemotherapy-related temporary disability). B-FED coordinates this care with Mom’s Meals Nourish Care (Ankeny, Iowa, www.momsmeals.com), a home delivery service that provides medically tailored meals across the U.S. Enrollment in Mom’s Meals requires a dietitian referral for the weekly number of meals delivered (ranges from 7-21) and type of diet. Before enrolling a patient in meal delivery, the B-FED Education Specialist conducts a home visit. The home visit aims to assess adequate storage capacity for the meals, verify a functioning refrigerator/freezer are available, and identify barriers to consuming prepared meals.

The B-FED Education Specialist communicates with the patient to discuss meal preferences and any dislikes or allergies before placing the order. Mom’s Meals calls the patient prior to the delivery of the meals to confirm date of delivery of the meals and to welcome them to the program. The meals are tailored to specific dietary requirements, ranging from general wellness to renal or low sodium. The patient receives a menu in the first delivery and can select meals from the tailored menu to facilitate patient self-empowerment and ownership of the process.

#### Food Preparation Items

Prior to B-FED, many BAO food assistance program participants reported lacking food preparation items necessary to safely prepare food in the home. Distribution of needed food preparation items began in tandem with the food box delivery, including can openers, measuring cups, cutting boards, meat thermometers and other items needed to prepare food received through the food box program. In addition, carrier carts are provided to patients relying on the public transit system or medical transport to assist in transporting the B-FED food box from BAO to the patient’s home. As some patients do not have running water and report obtaining water through other means (e.g., neighbor’s hose; rainwater and runoff), the B-FED Education Specialist verifies the patient’s access to clean, potable water. If water quality is a significant concern, a filtered-water pitcher with 12 months’ filter supply is provided throughout the duration of their enrollment in the program.

By providing items required for at-home meal preparation, the program is eliminating basic barriers to food consumption. For instance, if there is not a microwave to reheat home delivered frozen meals, or a patient can only consume pureed foods but has no blender, nutritional materials alone do not satisfy the end goal of positive food-driven health outcomes. Providing proper tools to reheat and cook items also aids in preventing foodborne illnesses.

#### Education

B-FED’s educational component focuses on food preparation, food safety, and nutrition. The overall aim of the education component is to reinforce the healthy grocery options provided in the food box and give patients confidence in using the provided food items. A phased program roll-out took place to generate interest and assess quality. Phase one consisted of “lunch and learns” on practical topics such as reading nutrition labels, basic nutrition, and interpreting lab results. A newsletter was included with all food boxes, discussing food safety, nutrition, and food preparation strategies for items in the food box. Phase two was an abbreviated course on food safety for PHW, which now serves as the introduction to the cooking class series. Cooking classes designed around monthly food box contents were developed to reinforce use of food box contents. Typically, 10 participants meet twice a month. To generate awareness and class participation, flyers with class information are on display throughout BAO and the 1917

Clinic for patients to see when patients collect monthly food boxes or attend other appointments. The monthly newsletter also includes class information. After each class, participants provide feedback via paper questionnaires to inform future classes. As interest in cooking classes grew, additional cooking class levels were implemented. In the spring of 2020, education workshops (centered around themes such as goal setting, stress management, etc.), support groups and cooking classes based on skillset and participation in prior courses were implemented. During the COVID-19 pandemic this series transitioned to an online Facebook and YouTube channel.

### Adjusting Services During the COVID-19 Pandemic

The B-FED program provided regular in-person contact from 2017-2020. Due to COVID-19 and concerns about compromising the health of patients, BAO altered food pick-up processes to a curb-side model in order to limit COVID-19 transmission risk for patients and BAO staff (Kay & Musgrove, 2020). Patient engagement remained high, with 79% of patient appointments completed between January and July of 2020. Additionally, the B-FED team adjusted food box components to maintain nutrition and food quality while accounting for temporary beef and pork product shortages, and price fluctuations in meat/poultry, eggs, cereals, and vegetables (Bureau of Economics).

## Results

Socio-demographic characteristics of the 1,055 B-FED program participants mirror the 1917 Clinic population with the majority of patients identifying as single, older, and Black race (**Table 2**). The B-FED program has a higher number of patients supported through Ryan White funding (96.1%) compared to the 1917 clinic overall population (80.1%). Initially, B-FED assisted an estimated 280 patients monthly. The program now assists over 700 patients per month with over 1,000 patients participating since the start of the program. The food program has reengaged more than 300 PWH who were previously lost-to-follow-up to re-engage in HIV care, which is critical to achieve viral suppression. Of those who returned to care, 30% returned after ≥ 5 years and 14% after ≥ 10 years of being out of care (**Figure 1**). When asked about their experiences with B-FED food assistance and education programs, patients provided examples of improved food quality (*“Y’all tell me to have some vegetables, I can do that now. And they good ones, too*.*”* – male in their 50s), ART adherence (*“They said I had to take my medicine with 300 calories; if I don’t got 300 calories I can’t take it. I don’t miss my dose now.”* – female in their 30s), and culinary skills (*“My daughter was telling me that my cooking classes are good for me because now my food tastes better.”* –female cooking class student).

**Table 2.**
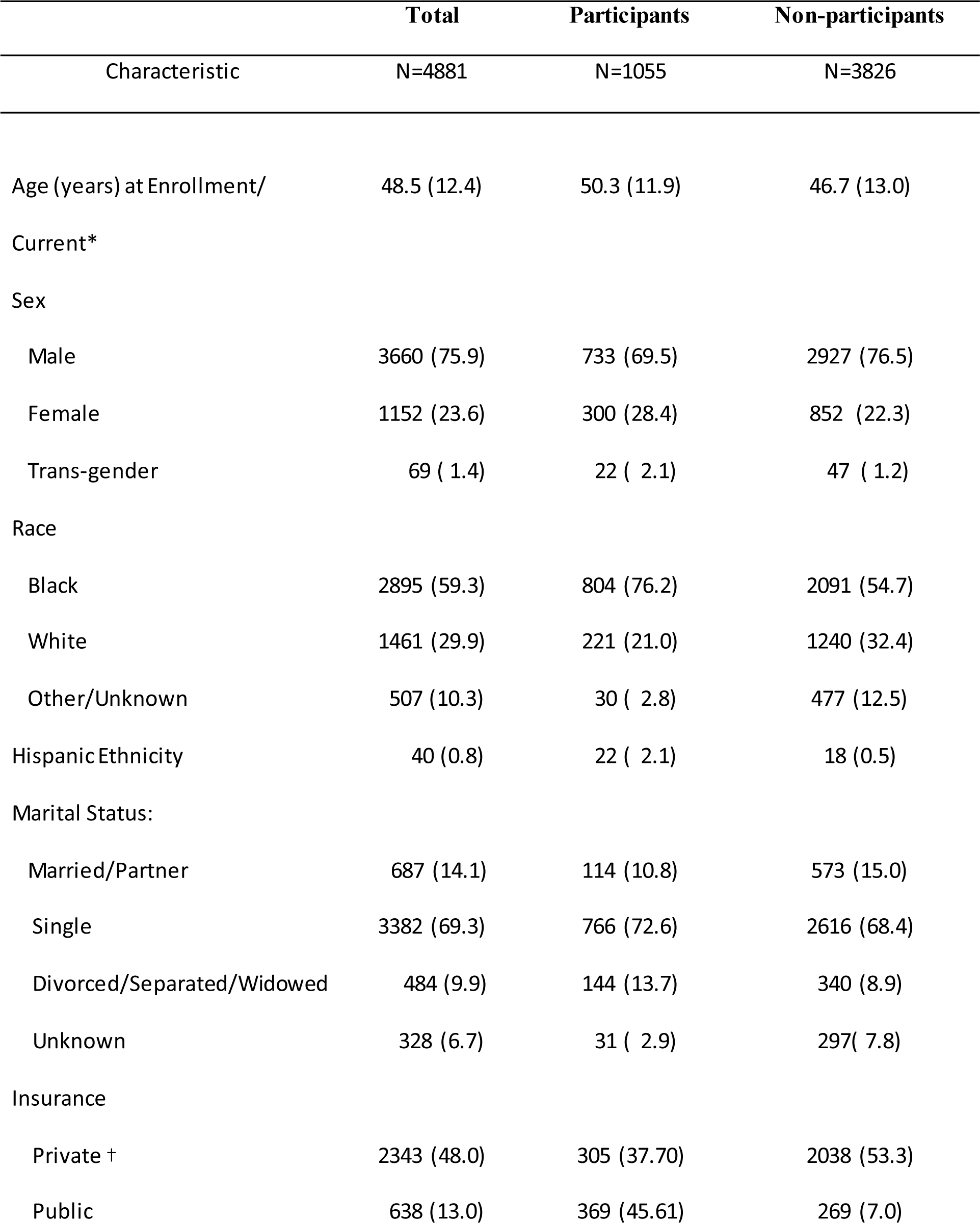
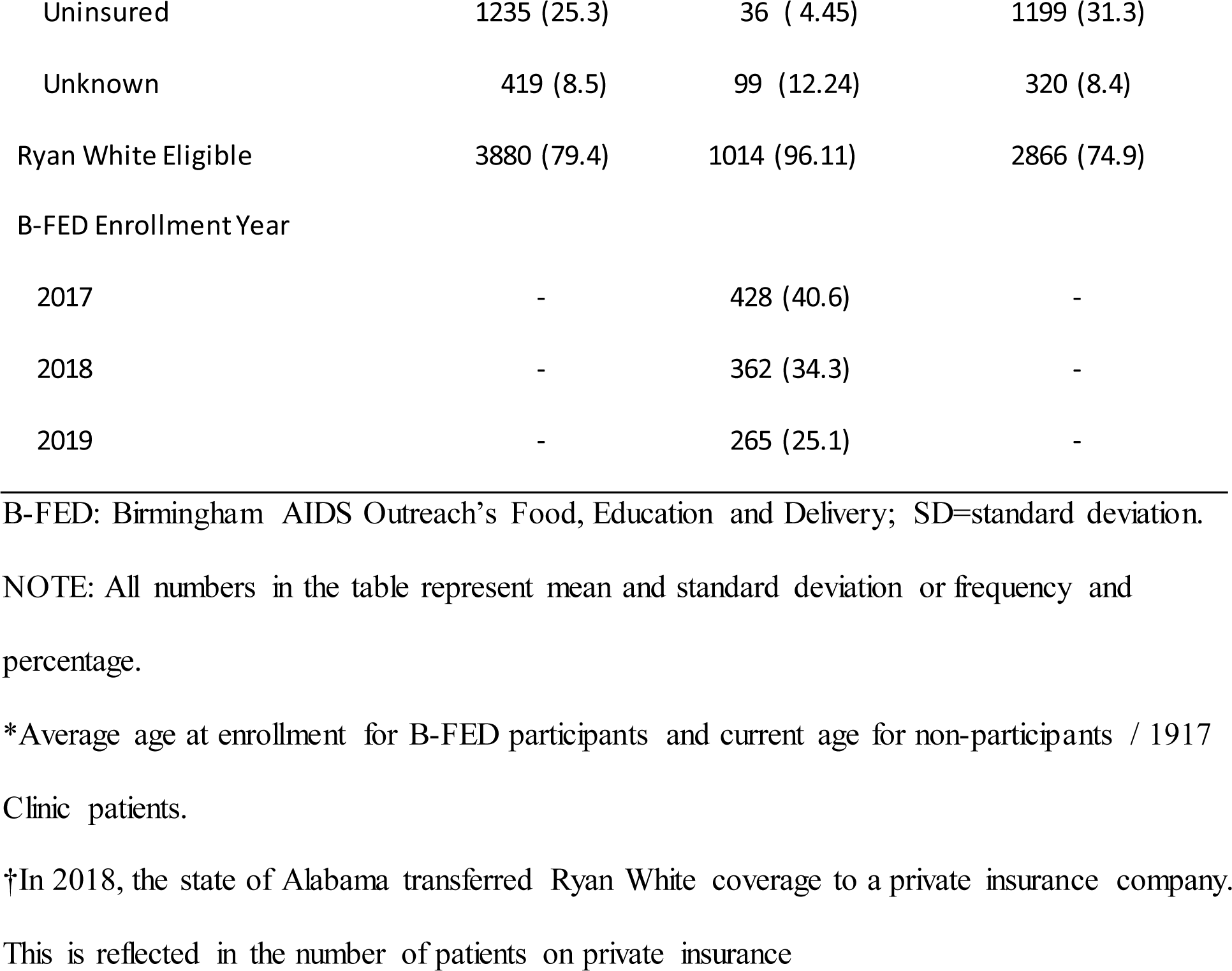
Socio-demographic characteristics of the 1917 Clinic population, B-FED program participants, and non-participants, 2017-2019.

**Figure 1.**
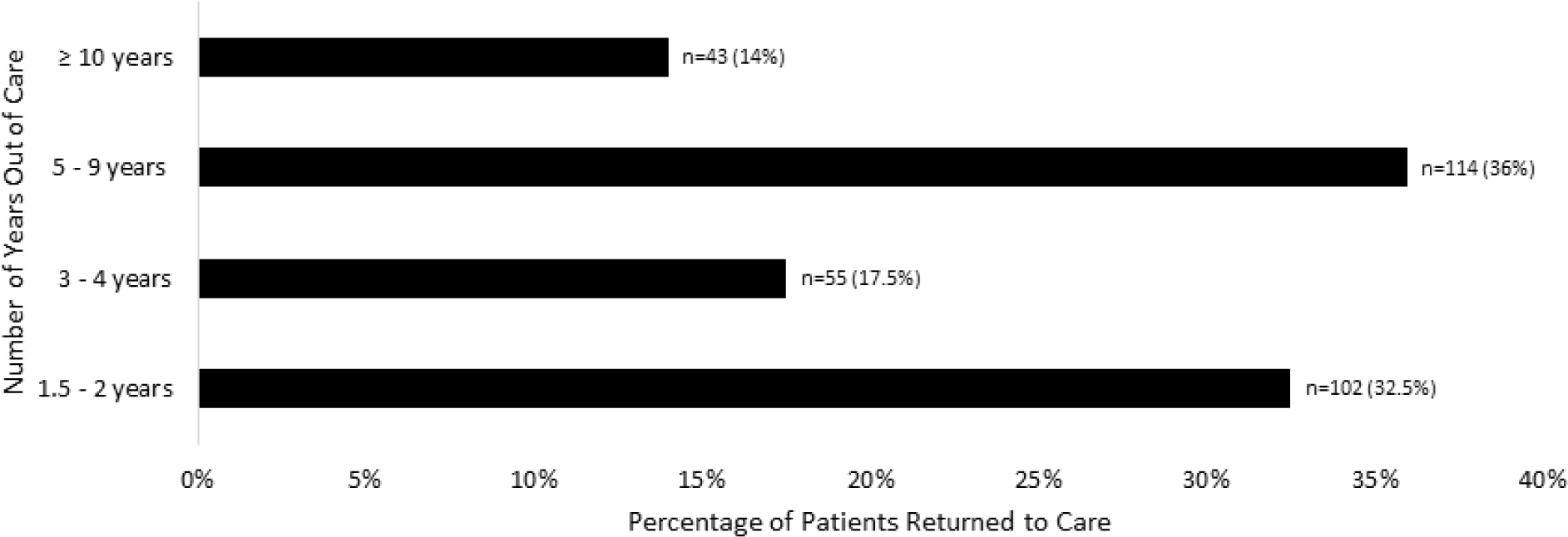
Length of time out of care for people with HIV (n=314) who re-engaged in HIV care through the B-FED program, 2017-2019.

## Discussion

The B-FED program has grown into a clinic/community partnership that uses a holistic approach to empower PWH to take control of their overall health. B-FED integrates patient feedback and program evaluations to better serve PWH in Birmingham, Alabama, and surrounding areas. The goal of future B-FED program evaluations is to report on the long-term impacts of the food assistance program on HIV treatment outcomes, quality of life, and metabolic health of participants. This novel clinic/community partnership to provide nutritionally adequate food assistance for PWH can serve as a roadmap for organizations to develop and tailor collaborative, sustainable food assistance programs to meet the food, medical, and social needs of adults with food insecurity, and improve clinical care and health outcomes for a variety of medical conditions.

## Supporting information

SQUIRE 2.0 Checklist

## Data Availability

All data produced in the present study are available upon reasonable request to the authors

## Acknowledgements

This implementation and quality improvement work was supported by 340B Program Income earned under the U.S. Department of Health and Human Services’ Human Health Resources and Services Administration Ryan White Part C Grant #H76HA25710. (PI: James Raper). ALW and DML were also supported by Award Number P30-AI27767 from the National Institute of Allergy and Infectious Disease, and Award Number P30-DK056336 from the National Institute of Diabetes and Digestive and Kidney Diseases. The authors would like to thank the patients, clients, care providers, and financial administrators at the 1917 Clinic and Birmingham AIDS Outreach for their thoughtful insights used to develop and monitor the B-FED program.

## Key Considerations

- Multi-faceted food assistance programs can serve as a key component to meet HIV care goals.
- Clear communication between clinic and community partners should be a priority for any food assistance program.
- Food assistance programs for PWH should consider needs beyond the basic provision of food, such as availability of safe water sources, necessary food preparation and storage equipment, and capacity to prepare own food versus receiving meal delivery.

