## Supplementary material for "Brief: Implementation of a Novel Clinic/Community Partnership Addressing Food Insecurity Among Adults with HIV in the Southern United States": SQUIRE 2.0 Checklist

| Section and Item | Page Number |
|---|---|
| **Title and Abstract** | |
| 1. Title | 1 |
| 2. Abstract | 3 |
| **Introduction** | |
| 3. Problem Description | 4 |
| 4. Available Knowledge | 4-5 |
| 5. Rationale | 5 |
| 6. Specific Aims | 4-5 |
| **Methods** | |
| 7. Context | 5-7 |
| 8. Intervention(s) | 7-12 |
| 9. Study of the Intervention(s) | 7-13 |
| 10. Measures | 12-13 |
| 11. Analysis | 12-13 |
| 12. Ethical Considerations | 7-8 |
| **Results** | |
| 13. Results | 12-13 |
| **Discussion** | |
| 14. Summary | 13 |
| 15. Interpretation | 13 |
| 16. Limitations | 13 |
| 17. Conclusions | 13, 16 |
| **Other Information** | |
| 18. Funding | 2 |
